# The Power of Partnership: Democratizing Genetic Prevalence to Empower Patient Advocacy

**DOI:** 10.64898/2026.03.30.26349539

**Authors:** Samantha M. Baxter, Moriel Singer-Berk, Carmen Glaze, Kathryn Russell, Riley H. Grant, Emily Groopman, Josephine Lee, Nick Watts, Jordan C. Wood, Michael Wilson, Rare As One Network, Heidi L. Rehm, Anne O’Donnell-Luria

**Affiliations:** Broad Institute of MIT and Harvard, Cambridge, MA, United States; Massachusetts General Hospital, Boston, MA, United States; Boston Children’s Hospital, Boston, MA, United States

## Abstract

**Introduction:** Accurate estimation of disease prevalence is crucial for public health and therapeutic development, but traditional methods are often inaccurate. Genetic prevalence, which estimates the proportion of a population with a causal genotype, using allele frequencies from population data, offers an important alternative.

**Methods:** We partnered with 18 Rare As One patient organizations to estimate genetic prevalence for 22 autosomal recessive conditions using population data from two releases of the Genome Aggregation Database (gnomAD). To standardize and democratize these analyses, we developed the Genetic Prevalence Estimator (GeniE), a publicly available tool, for accessible calculations.

**Results:** Conservative carrier frequencies in gnomAD v4.1 ranged from 1/164 to 1/11,888. The median change in genetic prevalence frequency between v2.1 to v4.1 was 0.806. Partnership with patient advocacy groups provided critical real-world context that refined the interpretation of these estimates.

**Discussion:** These findings highlight that genetic prevalence is not a static figure but a dynamic, evolving measure with important caveats that need to be considered. Our study underscores the necessity of re-evaluations as databases expand. By integrating patient-partnered insights with the GeniE platform, we empower the genomics community to maintain transparent, up-to-date, and actionable data for rare disease advocacy and drug development.

## Introduction

Accurate knowledge of disease prevalence guides decision-making across the healthcare system, including resource allocation for research, public health screening, and therapeutic development.^1^ Understanding prevalence both globally and within specific genetic ancestry groups (e.g., Admixed American, East Asian) informs advocacy efforts, facilitating outreach and network-building among affected individuals and families. Additionally, the carrier rate of recessive conditions is often used for deciding which genes are included in carrier screening panels.^2^ Yet, despite its importance, the carrier frequency and prevalence of most rare conditions remains unknown.

Traditional methods, such as case counting in or across health systems using ICD-10 codes, often yield biased and inaccurate estimates. Many rare conditions lack specific ICD-10 codes, and even when available, recognition by a physician or molecular diagnosis followed by proper coding are required for inclusion in electronic medical records. Newborn screening (NBS) is another method for estimating incidence, but NBS is only available for a small number of conditions, varies by state in the US and is incompletely implemented in many countries. Given these limitations, genetic prevalence, the estimated proportion of a population that has a causal genotype for a genetic condition, has become an increasingly utilized approach.^3^

Genetic prevalence for highly penetrant recessive conditions uses the presence of carriers to calculate the expected number of affected individuals by Hardy-Weinberg equilibrium (HWE).^4^ For a single disease-causing variant inherited in an autosomal recessive manner, the individual variant frequency can be doubled for carrier frequency or squared for genetic prevalence. For an autosomal recessive condition caused by a single gene, the same method can be used, but instead of using a single allele frequency, the method uses the sum of allele frequencies for all disease-causing variants in that gene before doubling (carrier frequency) or squaring (genetic prevalence). Despite the dynamic nature of variant classification and population allele frequencies, the vast majority of genetic prevalence estimates are static, using the variant classifications and allele frequencies available at the time of publication.^5^ Given the rapid pace of variant-level data sharing, genetic prevalence should be considered an evolving estimate, which is reassessed as new gene-level and variant-level information is learned.

Most of these estimates have been performed in an academic setting, with limited to no engagement with the relevant patient advocacy group(s). These groups not only have the most pressing need for this information, but they also provide critical insights on how these estimates do or do not align with real world data, such as patient registries. The lack of collaboration represents a missed opportunity to integrate genetic prevalence estimates with other methods of estimating condition burden.

In this manuscript, we describe our collaborative approach to estimating genetic prevalence for autosomal recessive (AR) monogenic conditions. We introduce the Genetic Prevalence Estimator (GeniE; broad.io/genie), a tool developed based on insights from this study.

Additionally, we propose best practices and tools for developing, sharing, and interpreting genetic prevalence estimates, ensuring greater accuracy and utility for researchers, industry and patient communities.

## Methods

### Collaboration with Patient Organizations

Since January 2021, our team has partnered with the Chan Zuckerberg Initiative’s Rare As One (RAO) network of patient organizations to estimate the prevalence of rare AR conditions.^6^ RAO patient organizations, along with their nexus of condition and gene experts, provided input throughout the study on: 1) gene selection, 2) additional sources of variation (e.g., internal lists, condition-specific databases), and 3) evidence towards pathogenicity (e.g., functional assays, expert review of evidence). Carrier frequency and genetic prevalence results were returned directly to the patient organizations and their experts via 90-minute video conferences, accompanied by written materials.

### Gene Selection

RAO patient organizations, with at least 1 known gene associated with an AR condition, were encouraged to participate by submitting gene nominations for the Genetic Prevalence Study. Some of the groups with multiple genes had two genes nominated. After carefully reviewing all submitted genes based on factors such as inheritance pattern, gene-condition validity, and available variant data, we selected 22 genes for inclusion.

### Variant Selection

For each gene included in this study, we collected all variants in the Genome Aggregation Database (gnomAD)^7^ that passed gnomAD’s QC filters (PASS), with an allele count (AC) of 1 or more, in the MANE Select or, where unavailable, in the Ensembl canonical transcript. Any variants that were flagged in gnomAD (e.g., AS VSQR, AC0) as suspected artifacts or sequencing errors were excluded from this study. We then filtered these to variants that met one or more of these criteria (Figure 1):

**Figure 1.**
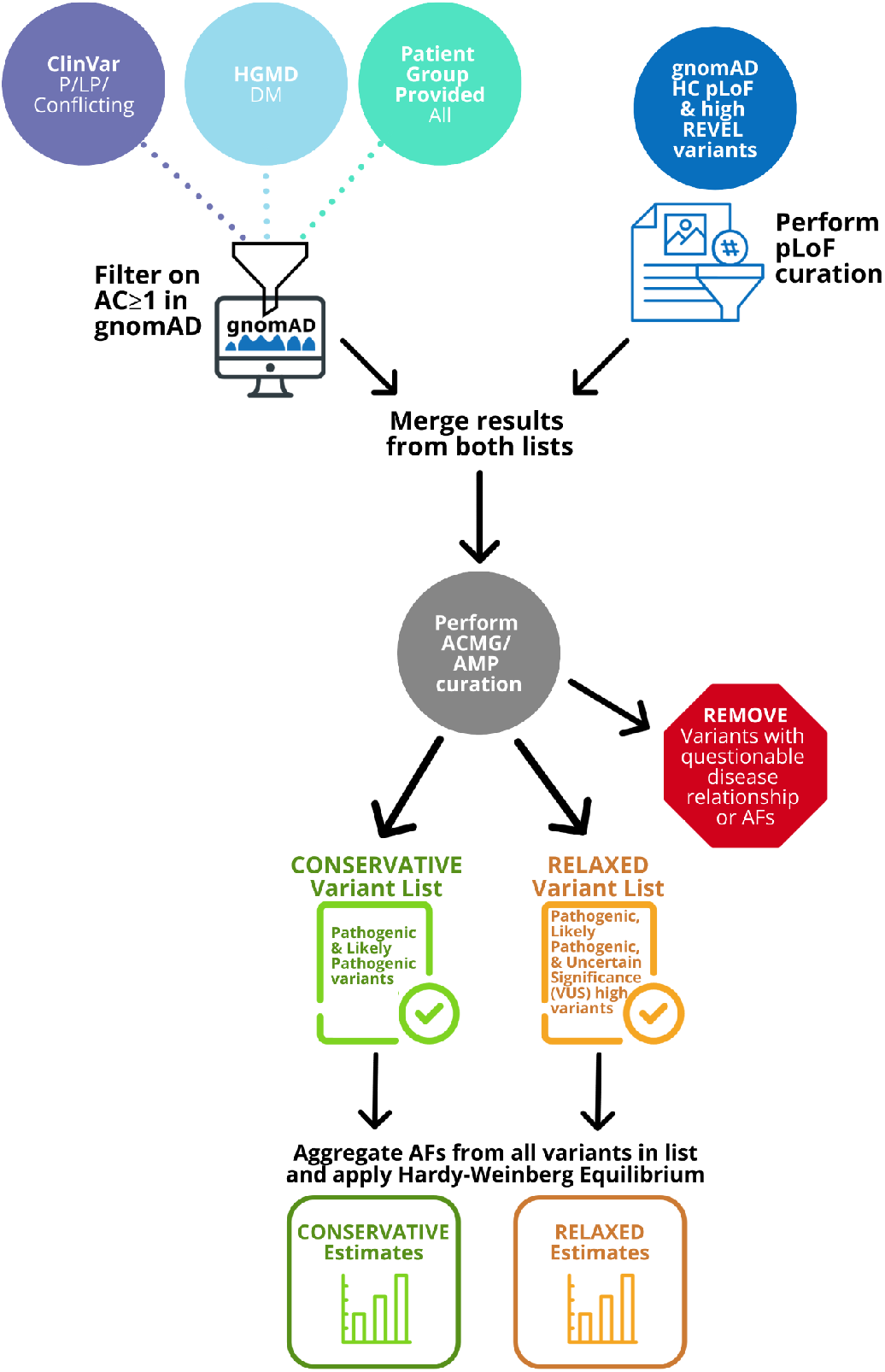
Workflow for the genetics prevalence study.

- <u>ClinVar</u>: All pathogenic (P), likely pathogenic (LP), and conflicting variants, where one of the conflicts was a P or LP submission.
- <u>HGMD</u>: All Disease Mutations (DM).
- <u>gnomAD</u>: High-confidence (HC) predicted loss of function (pLoF) variants and missense variants with a REVEL score ≥0.932^8,9^ (v4.1 only).
- <u>Other</u>: All candidate disease-causing variants provided by the patient organization, which were not previously included through the other sources.

### gnomAD version

When this collaboration started in 2021, gnomAD v2 was used in the project as it was the largest open-source population database available.^10^ When gnomAD v4.1 was released in April of 2024^11^, we reran variant selection using the updated data. Due to the large number of new variants from v4.1 that met our selection criteria across the 22 genes (n=4,600), we developed a triaged approach to identify and curate the variants that would have the most impact on the final estimate. All variants with an AC≥15 (allele frequency [AF] ≥ 1.0E-05) in v4.1 received a full curation, all previously reviewed v2 VUS, regardless of v4.1 AC, underwent an updated literature search to look for new evidence, and all new HC pLoF with an AC≥2 underwent pLoF curation.^12^ The ClinVar, HGMD or HC pLoF variants that were new in v4.1 underwent a thorough literature search and unless contradictory evidence was found, they were assumed to be at least LP and included in the conservative estimates. The one exception was gnomAD high REVEL-only variants, which were considered as VUS-high. This triage approach notably reduced the amount of curation time, while still ensuring that groups received the most up-to-date results. Having v2 and v4 results allowed us to assess the impact of gnomAD v4.1 on genetic prevalence, including changes in variant numbers, allele frequencies, and whether the estimates changed, both for the entire dataset, as well as for specific genetic ancestry groups.

### Variant Curation

All variants that passed gnomAD QC filters and met our criteria for inclusion were curated and classified using current variant classification guidelines.^13^ All gnomAD HC pLoF variants (defined as “stop_gained”, “frameshift_variant”, “splice_acceptor_variant”, or “splice_donor_variant” in the relevant transcript) underwent a specialized pLoF curation protocol to identify variants that did not result in LoF by nonsense mediated decay due to factors including but not limited to rescue.^12^ These pLoF curation results influenced the weighted strength of the PVS1 criteria.

If robust functional data were available for a gene, we integrated it into our approach. For example, in collaboration with the DADA2 Foundation, we combined our pLoF curation results with a list of functionally modeled *ADA2* variants demonstrating <25% residual enzyme activity, which is consistent with the threshold for phenotypic expression. Since this method was an outlier and not comparable to the method used for the other RAO genes, we did not include these previously published results^14^ in the summary section below. When applicable, other frameworks were used such as any Clinical Genome Resource (ClinGen) Expert Panel gene-specific guidelines or the ACMG and ClinGen copy number variant recommendations.^15^ All variants were classified using the ACMG/AMP^13^ five-tier system: Pathogenic (P), Likely Pathogenic (LP), Uncertain Significance (VUS), Likely Benign (LB), and Benign (B) (Supplemental File 1).

### Exclusion from Calculations

Before calculations were started, study staff reviewed curation results to determine if any variants should be excluded. Variants were excluded if they met one or more criteria 1) Classified as VUS, with limited/no case data, and an AF higher than the most common P/LP variant, 2) Classified as VUS-low, LB, or B, 3) Pathogenicity is not associated to the condition of interest, 4) Likely mapping errors, 5) pLoF variants in homopolymer regions which are enriched for sequencing artifacts, with insufficient evidence to support inclusion, 6) pLoF variants with multi-nucleotide (MNV) or frame-restoring events without additional pathogenicity evidence. All variants removed from analysis were discussed with the patient organization and their network for review and approval of removal.

### Calculations

Carrier frequency and genetic prevalence were estimated across all gnomAD samples, and for any gnomAD genetic ancestry group with >2,000 alleles.^10^ Estimates are calculated using gnomAD AFs and the Hardy-Weinberg equation, where:

q = variant AF

q_a_= aggregate AF

2q_a_= cumulative carrier frequency

q_a_^2^ = genetic prevalence

The “conservative” estimates included only P and LP variants, while the “relaxed” estimates also included VUS-high variants, which are a subclass of VUS that have some evidence to support pathogenicity but not enough to officially classify as LP.^12^ The ability to perform these calculations was developed into a web-based, open-source tool called the Genetic Prevalence Estimator (GeniE).

### Data Sharing

Variant curations completed in this project were submitted to ClinVar to increase the number of publicly available classified variants for these genes, which also helps improve the diagnostic accuracy for these conditions. ClinVar annotations and curations of the loss-of-function variants in gnomAD are displayed as annotations in the gnomAD browser on the gene and variant pages and can be downloaded from the data page.

### Genetic Prevalence Estimator (GeniE)

Current bioinformatic approaches to genetic prevalence estimation^2^ have pipelines that require individuals to have computational skills in order to use the tools and rely heavily on database-specific annotations to produce the estimates and require users to have computational skills. To standardize and increase the accessibility of genetic prevalence estimation, we created the Genetic Prevalence Estimator (GeniE; broad.io/genie), a publicly available tool that uses AFs from gnomAD (Figure 2). The tool was developed in collaboration with a community of experts and users, including patient advocacy groups. GeniE assists users in creating a list of known disease-causing variants and performing genetic prevalence calculations. It provides carrier frequency and prevalence results within minutes, which can be downloaded or shared directly with other users via the interface. Users can choose to make their lists and any supporting documentation public, allowing others to access and review their estimates. The variant data (including nomenclature, ClinVar classification, and gnomAD AF data) is provided for every variant, ensuring full transparency into how and when these estimates were created.

**Figure 2.**
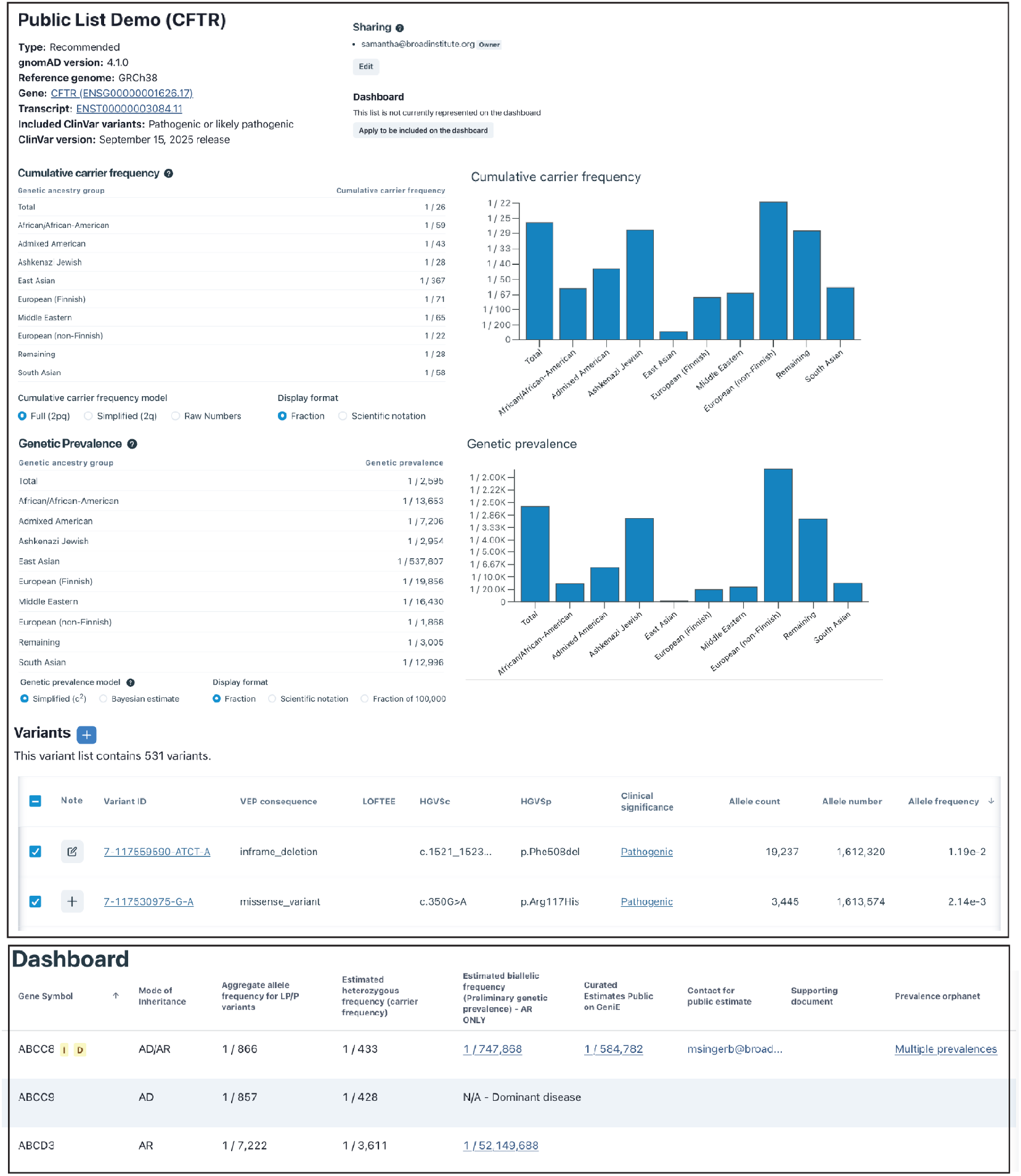
Screenshots of GeniE. (Top panel) Genetic prevalence calculator (Bottom panel) GeniE Dashboard

In addition to the calculator, we developed the GeniE Dashboard, which serves as a lookup tool for preliminary genetic prevalence estimates and a landing page for public GeniE lists. These preliminary estimates are based on gnomAD v4.1 frequencies for ClinVar LP/P variants and gnomAD HC pLoF variants. The top 10 highest frequency variants are displayed on the browser for quick review. However, as these lists are not manually curated, they should be used with caution as they may contain non-disease-causing variants. Orphanet prevalence estimates^16^ are also imported to the dashboard, for comparison, through their release files.

## Results

### Genetic Prevalence Estimates

Conservative carrier frequencies in gnomAD v4.1 for all the samples in gnomAD ranged from 1/164 to 1/11,888, with genetic prevalence estimates between 1/194,753 and 1/94,984,678 (Table 1). Relaxed estimates showed carrier frequencies from 1/140 to 1/1,101 and a prevalence range from 1/107,234 to 1/565,277,193 (supplemental table 1). The median prevalence ratio in conservative genetic prevalence from v2.1 to v4.1 was 0.806, with a wide range of 0.50 to 14.96-fold. Changes were primarily driven by updated AF or the presence of new variants. New variants encompass both those that were not present in gnomAD v2 but are now in v4, and variants that had an AC≥1 in v2, but did not meet other criteria for inclusion at the time. Notably, 1,633 (81.0%) of the 2,016 variants present in both gnomAD v2 and v4 had a decreased AF in v4.1, consistent with the increased resolution of AF expected from a larger reference database, particularly for ultra rare variants. All variants used in these calculations, along with classifications, v2 AF, and v4 AF are included in supplemental table 2. The wide range of prevalence estimates and the decrease in AF for most variants highlight the importance of regularly updating these estimates as variant classifications and population databases evolve.

**Table 1.** Summary of v2.1 and v4 .1 conservative results.

| Gene | v2.1 CF | v4.1 CF | v2.1 GP | v4.1 GP | v4.1 GeniE Link | Prevalence Ratio (GP) | Primary variants impacting change in CF and GP |
| --- | --- | --- | --- | --- | --- | --- | --- |
| *ABCB11* | 1/429 | 1/525 | 1/734,727 | 1/1,103,934 | broad.io/GeniE/ABCB11 | 0.666 | 9 out of the top 10 v2 variants have reduced AF in v4.1. |
| *ABCC8* | 1/318 | 1/382 | 1/404,541 | 1/584,782 | broad.io/GeniE/ABCC8 | 0.692 | Top 10 v2 variants have reduced AF in v4.1. |
| *DARS2*<sup>a</sup> | 1/510 | 1/616 | 1/1,096,513 | 1/1,611,618 | broad.io/GeniE/DARS2 | 0.680 | 8 out of the top 10 v2 variants have reduced AF in v4.1. |
| *DNAH11* | 1/168 | 1/236 | 1/112,422 | 1/223,101 | broad.io/GeniE/DNAH11 | 0.504 | Cumulative impact of >82% of v2 *DNAH11* variants had reduced AF in v4.1. |
| *EFL1* | 1/8078 | 1/10613 | 1/260,986,896 | 1/45,0581,951 | broad.io/GeniE/EFL1 | 0.579 | All v2 variants have reduced AF in v4.1. |
| *EPM2A* | 1/1150 | 1/1557 | 1/5,292,985 | 1/9,697,019 | broad.io/GeniE/EPM2A | 0.546 | All but 2 of the v2 LP/P variants have reduced AF in v4.1. |
| *GAMT* | 1/990 | 1/464 | 1/3,923,670 | 1/860,867 | broad.io/GeniE/GAMT | 4.558 | The 4 most common v2 pathogenic variants had increased AF in v4.1. |
| *GBE1* | 1/284 | 1/243 | 1/323,188 | 1/235,784 | broad.io/GeniE/GBE1 | 1.371 | The most common v2 pathogenic variant (c.691+2T>C) had an increased AF in v4.1. Another VUS v2 variant was reclassified to LP (p.Glu609Lys). |
| *HADH* | 1/11888 | 1/4994 | 1/565,277,193 | 1/99,758,419 | broad.io/GeniE/ | 5.666 | A VUS v2 variant was reclassified to LP |
|  |  |  |  |  | HADH |  | (p.Gly303Ser) and is now the most common LP/P variant in v4.1. |
| *HPS1* | 1/585 | 1/651 | 1/1,367,366 | 1/1,697,270 | broad.io/GeniE_HPS1 | 0.806 | Cumulative impact of >83% of *HPS1* v2 variants had reduced AF in v4.1. |
| *HPS4* | 1/2147 | 1/2912 | 1/18,434,176 | 1/33,914,574 | broad.io/GeniE_HPS4 | 0.544 | Cumulative impact of >90% of *HPS4* v2 variants had reduced AF in v4.1. |
| *NEB* | 1/164 | 1/221 | 1/107,234 | 1/194,753 | broad.io/GeniE_NEB | 0.551 | 19 out of the top 20 v2 variants have reduced AF in v4.1. |
| *PCDH15* | 1/522 | 1/656 | 1/1,090,265 | 1/1,720,051 | broad.io/GeniE_PCDH15 | 0.634 | All top 15 variants have reduced AF in v4.1. |
| *PLA2G6* | 1/691 | 1/493 | 1/1,912,273 | 1/970,522 | broad.io/GeniE_PLA2G6 | 1.970 | The top 5 variants in v4.1 had an increased AF from v2 or was a new LP/P variant. |
| *POLR3A\** | 1/228 | 1/156 | 1/207,677 | 1/226,819 | broad.io/GeniE_POLR3A | 0.916 | Top variant (c.1909+22G>A) increased AF from v2 and v4.1, but homozygotes were removed in v4.1 estimate due to new evidence that it only causes disease in the compound heterozygous state. |
| *POLR3B* | 1/526 | 1/251 | 1/1,105,309 | 1/251,628 | broad.io/GeniE_POLR3B | 4.393 | The most common v2 pathogenic variant (p.Val523Glu) had an increased AF in v2.1. A v2 VUS variant was reclassified to LP (p.Met415Thr), and is now the most common variant in v4.1. |
| *POMGnT1* | 1/436 | 1/533 | 1/761,697 | 1/1,136,560 | broad.io/GeniE_POMGnT1 | 0.670 | The most common LP/P variant (c.1539+1G>A) had a reduced AF between v2.1 and v4.1. |
| *SELENON* | 1/460 | 1/387 | 1/845,135 | 1/598,419 | broad.io/GeniE_SELENON | 1.412 | 3 out of top 4 v2 LP/P variants, including most common (p.Gly315Ser), had increased frequency. Addition of a new LP variant that is now 5th most common in v4.1 (c.1A>G; p.Met1?). |
| *SLC13A5* | 1/1856 | 1/1659 | 1/13,780,637 | 1/1,1006,411 | broad.io/GeniE_SLC13A5 | 1.252 | 3 out of top 6 v4.1 variants had an increased AF and a v2 VUS variant was reclassified to LP (p.Pro505Leu). |
| *TANGO2* | 1/564 | 1/614 | 1/1,270,473 | 1/1,509,686 | broad.io/GeniE_TANGO2 | 0.842 | Exon 3-9 deletion had slightly increased frequency, but one of the most common variants (p.Gly154Arg) decreased in frequency in v4.1. |
| *TBCK* | 1/1883 | 1/487 | 1/14,186,359 | 1/948,489 | broad.io/GeniE_TBCK | 14.957 | Top 3 v4.1 variants all had increased frequency, plus there are 2 new LP variants that are in the top 10 most frequent. |

CF= Carrier Frequency, GP= Genetic Prevalence
<sup>^</sup>For *DARS2* homozygosity and compound heterozygosity of variants predicated to cause nonsense mediated decay are believed to not be compatible with life. To address this we used the formula $q_a^2 - q_{nmd}^2$ , where $q_{nmd}$ = the aggregate allele frequency of all variants predicted to undergo nonsense mediated decay (nonsense and frameshift where LoF curation showed no evidence against NMD). See the supplemental case studies more details on these methods; \*For *POLR3A* c.1909+22G>A only leads to disease when seen in compound heterozygosity. We addressed this by using the formula $q_a^2 - q_{c.1909+22G>A}^2$ .

For some genes, genetic ancestry-group-specific carrier frequencies increased even when the cosmopolitan (e.g., estimates derived from all samples in gnomAD) carrier frequency and genetic prevalence decreased. For the *ABCC8* gene, a decrease in the cosmopolitan carrier frequency (prevalence ratio 0.69) was observed, yet the carrier frequencies for the African (AFR) and East Asian (EAS) ancestry groups increased by 2.0- and 3.4-fold, respectively. This was driven by the discovery of 35 new variants in the AFR group and 26 new variants in the EAS group. Similarly, for *HPS1*, the cosmopolitan carrier frequency decreased, but the Puerto Rican founder variant (p.His497fs) increased in frequency from v2.1 (AF = 3.19×10^−5^) to v4.1 (AF = 5.20×10^−5^), likely due to the inclusion of more Admixed American individuals in gnomAD v4.1 (v2 = 35,440; v4 = 60,038). All v4.1 genetic ancestry-group-specific carrier frequencies are available via the GeniE links in Table 1.

### Variant source comparison

Figure 3 illustrates the contribution of variants from different sources used to estimate the conservative carrier frequency. Out of the 6,616 v4 variants that met our inclusion criteria, approximately 68.4% (n=4,525) had an entry only in one source, 22.8% (n= 1,510) were found in 2 sources and 8.78% (n=581) were found in all three sources (supplemental table 3). To assess how many of these variants can be found in free public repositories, we broke down the carrier frequencies of each gene by source (Figure 3b) starting with ClinVar (85.6%), followed by gnomAD pLoFs not in ClinVar (11.8%) and ending with those only in HGMD (2.5%), a subscription-based commercial database. This highlights that the vast majority of disease-causing variants are represented in free, public databases including ClinVar and gnomAD.

**Figure 3.**
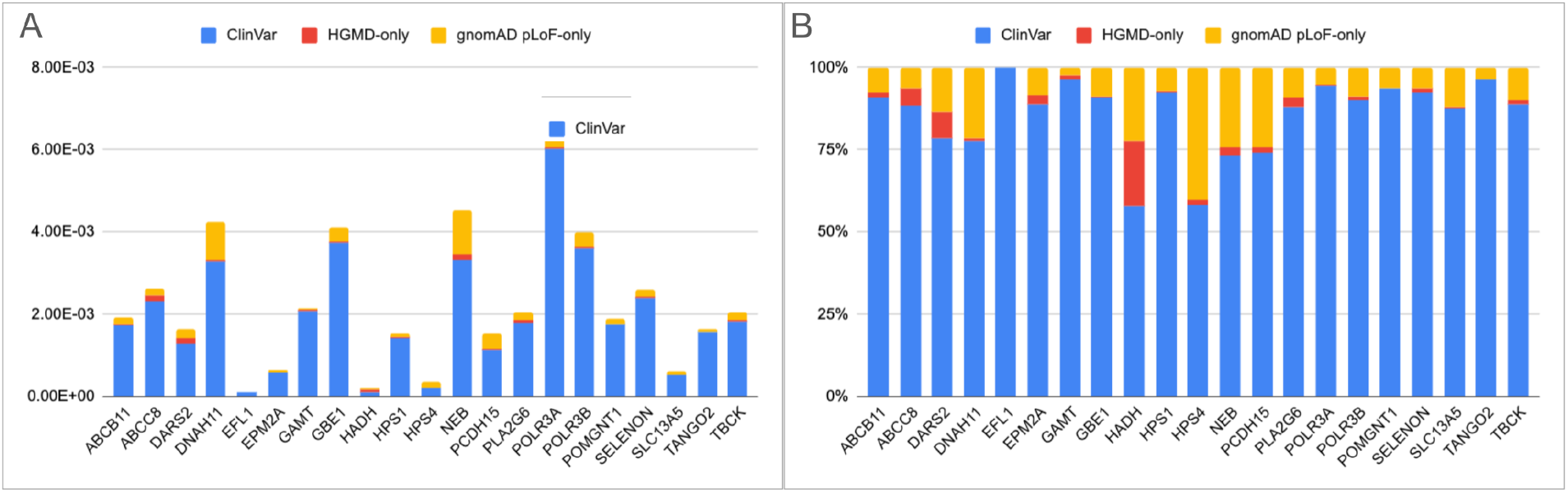
Contribution of source to v4 conservative carrier frequency. A) Contribution of source to total cumulative carrier frequency, B) Percentage contribution of source total cumulative carrier frequency

Our team performed curations on the 2,291 variants included in v2 as well as the 55 new v4 variants (7.7%) that had an AC ≥15. A comparison of our post-curation results with the pre-curation classification from various sources showed a 76.5% agreement with ClinVar’s pathogenic/likely pathogenic (P/LP) classifications, but only a 48.7% agreement with HGMD disease mutation (DM) classifications. Notably, for variants listed only in HGMD, the agreement rate went down to 18.7% (Table 2).

**Table 2.** Classification comparison.

| Initial classification vs study classification analysis |  | # of variants | Final Variant Classification |  |  |  |  |  |
| --- | --- | --- | --- | --- | --- | --- | --- | --- |
|  |  |  | Path/LP |  | VUS - High |  | Do Not Include (VUS or lower) |  |
| ClinVar | Path/LP | 757 | 579 | 76.49% | 151 | 19.95% | 27 | 3.57% |
|  | Conflicting | 154 | 44 | 28.57% | 88 | 57.14% | 22 | 14.29% |
| HGMD | DM (All) | 1051 | 503 | 47.86% | 393 | 37.39% | 155 | 14.75% |
|  | DM (HGMD-only) | 485 | 86 | 17.73% | 261 | 53.81% | 138 | 28.45% |

### Feedback from Organizations

In August 2025, we collected feedback from 17 RAO groups regarding their results and how they utilized the estimates in their work. Out of the 17 groups, 16 rated their estimates as “highly impactful,” while one group rated the impact as “moderately impactful.” When asked about expectations, 5 genes had lower estimates than expected, 5 were higher than expected, 4 were as expected, and 8 had no prior expectations. Groups with lower-than-expected estimates often had other data suggesting that this method might not fully capture all patients, perhaps due to variants being more common in underrepresented populations (e.g., *HADH*) or specific variant types not being available. *HADH* is reported to be more common in the Middle Eastern population, primarily due to a known Turkish founder variant,^17^ but also due to additional variants found in affected patients with known Middle Eastern descent (personal communication with Congenital Hyperinsulinism International and The Exeter Genomics Laboratory, UK). Due to the underrepresentation of this genetic ancestry group in gnomAD, none of these variants are present in the database, likely leading to an underestimate of carrier frequency and genetic prevalence. We propose various strategies for tackling different issues that may arise in the supplemental case studies. Patient organizations reported using this data for population outreach, presentations at patient and scientific conferences, communication with BioPharma companies, and manuscript submission.^14,18–20^ Gene and group-specific feedback is detailed in Table 3, and specific details of how each group has used this data in their work can be found in supplemental table 4.

**Table 3.** Organizational feedback on genetic prevalence estimates.

| Patient Organization | Gene | Feedback from Org. | Additional comments |
| --- | --- | --- | --- |
| A Cure for Ellie/Cure LBSL | *DARS2* | Higher than expected |  |
| A Foundation Building Strength | *NEB* | No prior expectations |  |
| APBD Research Foundation | *GBE1* | Higher than expected | Publication (https://www.medrxiv.org/content/10.64898/2025.12.16.25342386.v1). Used data as proof of concept for funding further natural history studies and justify therapeutic development |
| Association for Creatine Deficiencies | *GAMT* | Lower than expected | Ongoing collaboration with ACD and the ClinGen Cerebral Creatine Deficiency Syndromes Variant Curation Expert Panel (VCEP) to clarify classification of variants in ClinVar |
| Chelsea's Hope Lafora Children Research Fund | *EPM2A* | No prior expectations |  |
| Congenital Hyperinsulinism International | *HADH* | Lower than expected | Underestimate likely due to underrepresentation of Middle Eastern individuals in gnomAD. Known Turkish founder variant absent from gnomAD v2, as well as other variants previously identified in the middle eastern population. |
|  | *ABCC8* | Lower than expected | Work ongoing to find/add additional variants. Multiple conditions and inheritance patterns may also impact the accuracy of the results |
| Cure CMD | *SELENON* | No prior expectations |  |
|  | *POMGnT1* | No prior expectations |  |
| DADA2 Foundation | *ADA2* | Higher than expected | Publication (PMID: 34004258) |
| Hermansky Pudlak Syndrome Network | *HPS1* | As Expected | Estimates are likely an underestimate due to known Puerto Rican (PR) founder variant being lower than expected in v4, likely due to a low number of PR individuals in gnomAD. |
|  | *HPS4* | Higher than expected |  |
| INADcure Foundation | *PLA2G6* | As Expected | Publication (PMID: 39425167) |
| PCD Foundation | *DNAH11* | As Expected |  |
| Progressive Familial Intrahepatic Cholestasis Advocacy and Resource Network, Inc. | *ABCB11* | No prior expectations |  |
| Shwachman-Diamond Syndrome Alliance Inc | *EFL1* | No prior expectations | *SBDS* was not able to be analyzed due to pseudogene with high homology, *EFL1* is a far more rare cause with limited known pathogenic variants |
| TANGO2 Research Foundation | *TANGO2* | Lower than expected | Publication (PMID: 36473599); This estimate does not address the frequency of *TANGO2* deficiency in 22q11 deletion syndrome. See supplemental case studies for more detail.<br>The data showing that the Admixed American genetic ancestry group has a higher carrier frequency than the general population was used in a grant to justify providing more access for genetic testing in communities along the Mexico/Texas border. |
| TESS Research Foundation | *SLC13A5* | Lower than expected | Work ongoing to find/add additional variants and clarify classification of missense VUS |
| The TBCK Foundation | *TBCK* | Higher than expected |  |
| The Yaya Foundation for 4H Leukodystrophy | *POLR3A* | No prior expectations | Estimates confirmed concerns that POLR3-related Leukodystrophy may be underdiagnosed and are used for planning. |
|  | *POLR3B* | No prior expectations |  |
| Usher 1F Collaborative | *PCDH15* | As Expected | Numbers have been used for informing next steps in their strategic plan. |

## Discussion

Genetic prevalence is a powerful mechanism for estimating the number of individuals with a genetic condition, especially for rare and challenging-to-diagnose conditions. Unlike traditional prevalence and incidence metrics, which rely on symptomatic cases, genetic prevalence is based on the presence of disease-causing variation in an individual’s genome.^3^ This study highlights that genetic prevalence estimates that rely on regularly updated knowledge resources are dynamic and should not be considered in isolation of other data sources that inform prevalence. Estimates require ongoing refinement and monitoring over time. Including patient advocacy organizations in this process provides crucial context, additional data, and scientific and clinical expertise, which ensures a more accurate interpretation of the results.

### Key Elements Impacting the Results

Consistent with previous reviews,^5^ we identified five key factors that influence the interpretation of genetic prevalence estimates: 1) representation, 2) curation, 3) clinical sensitivity, 4) gene-disease novelty, and 5) disease spectrum.

1. *<u>Representation</u>*: Missing or underrepresented ancestry groups in population data can lead to either over- or underestimation of genetic prevalence. While gnomAD is one of the largest databases, it is not representative of the entire global population. Variants identified in underrepresented genetic ancestry groups or regions only detectable by genomes are more susceptible to sample bias.^21^ Some genetic ancestry-group-specific variants may be absent or present at very low frequencies, leading to an underestimation of the condition in that population. **The absence of any pathogenic variants in a specific genetic ancestry group should never be interpreted as an indication that the population is not at risk for a given condition**.
2. *<u>Curation of Variation</u>*: The foundation of these estimates is the list of variants included. Variants that do not cause the condition are under less selective pressure and can have higher AFs in the general population. The inclusion of even a single misclassified or hypomorphic variant can dramatically impact the results. **This analysis showed reasonable but not complete (76%) agreement with ClinVar classifications, emphasizing that investigators should perform a level of curation to ensure all included variants are genuinely associated with the condition of interest**.
3. *<u>Sequencing Technology Sensitivity:</u>* Furthermore, variants in gnomAD and other commonly used databases currently are typically identified via short read next generation sequencing. **Variants not detectable by these technologies used to generate the population data will not be included in these estimates, and variants only detectable by genomes may be underascertained given most of the data is from exome sequencing**.
4. *<u>Gene-Disease Novelty:</u>* Genes with more recent disease associations may have fewer established pathogenic variants, leading to a lower estimated prevalence. **The novelty of the gene-disease relationship, and the current number of reported pathogenic variants should be taken into context when assessing the accuracy of the estimation**.
5. *<u>Disease spectrum</u>*: The spectrum of a condition has a large influence on the interpretation of genetic prevalence estimates. The following aspects of a condition’s spectrum should always be considered.

<u>Symptomatic carriers</u>: This study uses gnomAD as the source of AFs. Individuals with some presentation/features of the condition are less likely to participate in or meet recruitment criteria for common disease research studies, which are the basis of this reference population database. **Depletion of symptomatic heterozygous carriers could lower the allele frequencies of pathogenic variants observed in gnomAD**.

<u>Impacts to life expectancy</u>: Genetic prevalence estimates do not account for the possibility that certain allele combinations may lead to early miscarriage, stillbirth, or shortened life expectancy. **These scenarios may lead to a genetic prevalence estimate that is higher than actual disease prevalence**.

<u>Reduced penetrance and variable expressivity</u>: The full phenotypic spectrum is still being discovered for many conditions. **This method as described above does not account for reduced penetrance or variable expressivity substantive enough to be considered a distinct disorder**.

We provide tips for navigating these considerations along with case examples and suggestions for alternative models to address many of these elements and others in the supplement (supplementary case studies).

### Additional considerations for these estimates

*Database updates*: Results should be revisited after new major releases of population databases. Note that this study used the secondary releases (e.g., gnomAD v2.1 and v4.1) that have resolved identified issues from the initial releases.

### Mosaicism or germline selection

This method only works for non-mosaic germline variation, and does not take germline selection into account. While this is typically more common in dominant conditions, it should still be considered when interpreting results for recessive conditions.

### Collaboration with Patient Advocacy Groups

A primary goal of this study was to demonstrate the utility of a strong collaboration with patient advocacy groups. These groups and their network of experts were critical to the interpretation of the results. Their feedback on variants, especially whether they had ever been reported in cases, known populations with higher frequencies, unpublished data and connections to clinical and scientific experts shaped our understanding of the validity of these estimates. Having their input early in the process allowed us to refine our estimates or perform additional follow-up, if needed. Their insights and honest feedback shaped the key factors of interpretation. By providing the results directly to their organizations, they were able to use the data to best serve their communities, whether for publications, engaging with biopharma companies, or informing recruitment strategies.

### Considerations and Limitations

Genetic prevalence has its own benefits and limitations, distinct from traditional prevalence and incidence, which should be taken into account when interpreting estimates. HWE has its own assumptions, including 1) no new (*de novo*) mutations, 2) random mating, 3) no natural selection, 4) large population size (to avoid genetic drift), and 5) no gene flow (no migration).^5^ These assumptions often do not hold in the real world, which does impact the interpretation and accuracy of results. Random mating also does not account for increased consanguinity rates in some cultures, which can increase the prevalence of recessive conditions in those regions.

While other methods have included confidence intervals, these typically only account for the size of the genetic ancestry group, overlooking crucial factors such as clinical sensitivity, impact on life expectancy, and penetrance of the condition. We felt that including confidence intervals would overemphasize one factor and mislead users.

### Future Directions

Currently, GeniE only returns results for fully penetrant AR conditions. We have multiple features on the roadmap that will allow users to account for variants with reduced penetrance, exclude specific non-disease-causing combinations of variants (e.g., homozygous hypomorphic alleles), and combine results from multiple genes into a single result for conditions with locus heterogeneity.

Currently, this method is designed for genes associated with conditions with AR inheritance and would require modification for autosomal dominant or X-linked conditions. Our team plans to adapt these methods for estimating prevalence of autosomal dominant conditions^22,23^ and incorporate them into GeniE, with modifications to address gene and disease-specific considerations. While some past studies have attempted to use this method for X-linked recessive conditions, it can lead to underestimates. This is likely due to two factors: 1) the method does not account for the variable contributions of *de novo* pathogenic variants across X-linked conditions^24^, and 2) we see a depletion of female carriers for X-linked conditions in gnomAD, likely due to females with milder symptoms of X-linked condition being less likely to participate in research studies included in gnomAD. More work is needed to improve the quality of genetic prevalence estimates for all inherited conditions.

With GeniE and careful consideration of population data, estimating genetic prevalence from population variant data is a powerful and accessible way to understand the global impact of a rare disease and to prioritize therapeutic development strategies appropriate for each condition.

### GeniE Code and Resources

Additional details on how to use GeniE, as well as links to FAQs and technical specifications, can be found at: https://gnomad.broadinstitute.org/news/2024-06-genie. The source code is available on GitHub (https://github.com/broadinstitute/genetic-prevalence-estimator).

## Supporting information

Supplemental Tips and Case Studies

## Data Availability

Variant classifications have been submitted to ClinVar. All genetic prevalence estimates are public on the Genetic Prevalence Estimator (GeniE) Dashboard. The source code for GeniE is available on GitHub.

https://genie.broadinstitute.org/dashboard/

https://www.ncbi.nlm.nih.gov/clinvar/

https://github.com/broadinstitute/genetic-prevalence-estimator

## Rare As One Network (Banner Authorship List)

H. Orhan Akman^1,2^, Rachel Alvarez^3^, Donna Appell^4^, Alan H Beggs^5,6,7^, Sarah Brengosz^8^, Tanya L. Brown^9,9^, Melissa K. Chaikof^10^, Eugene P. Chambers^11,12^, Deena L. Chisholm^13^, Erin Chown, MS^14^, David P. Corey^15^, Stacy A. Cossette^16^, Katherine Donohue^17^, Emily Durham^18,19^, Gustavo Dziewczapolski^3,20^, Sarah E. Flanagan^21^, Ann Geffen^13^, Bernadette R. Gochuico^22^, Deberah Goldman^14^, Jennifer L. Goldstein^23^, Valerie Greger^24,25^, Eszter Hars^26^, Amanda Hope^27^, Divya Jayaraman^28^, Rebecca L. Koch^29^, Seema R. Lalani^30^, Pui Y. Lee^15,28^, Jeff Levenson^14^, Qing Lu^2,31^, Samuel J. Mackenzie^32^, Michele Manion^33^, Michael McGinn^34^, Elizabeth McGinn^34^, Joshua K. Meisner^35^, Lindsay T. Michalovicz Gill^14^, Christina Y. Miyake^30,36^, Matthew Morgan^14,37^, Mike Morris^13^, Kasha Morris^13^, Christina Nemeth^38,39^, Kimberly L. Nye^9,9^, Jennifer Orthmann-Murphy^2,40^, Leena Panwala^27^, Tai LS Pasquini^41^, Natacha Pires^14^, Julie Raskin^41^, Harriet Saxe^14^, Adam J Shapiro^42^, Nikki Shearer^18^, Nikki Stusick^18^, Ashley S. Thompson^26,43^, Jen Tozer^20^, Emily Ventura^8^, Heidi Wallis^44^

^1^Columbia University, New York City, NY, USA

^2^APBD Research Foundation (Scientific and Medical Advisory Board), New York, NY, USA

^3^Cure CMD, Lakewood, CA, USA

^4^Hermansky-Pudlak Syndrome Network, NYHouston, Texas, USA

^5^Division of Genetics and Genomics, Boston Children’s Hospital, Boston Children’s Hospital, Boston, MA, USA

^6^The Manton Center for Orphan Disease Research, Boston Children’s Hospital, Boston, MA, USA

^7^Department of Pediatrics, Harvard Medical School

^8^PFIC Network, Clay City, KY, United States

^9^TESS Research Foundation, Menlo Park, CA, USA

^10^Usher 1F Collaborative, Newtonville, MA, USA

^11^DADA2 Foundation, Nashville, TN, USA

^12^Vanderbilt University Medical Center, Nashville, TN, USA

^13^TANGO2 Research Foundation, Hadlyme, CT, USA

^14^APBD Research Foundation, New York, NY, USA

^15^Harvard Medical School, Boston, MA, USA

^16^AFBS for Nemaline Myopathy, Palo Alto, CA. USA

^17^Chelsea’s Hope Lafora Children Research Foundation, Lexington, KY USA

^18^The TBCK Foundation, Pittsburgh, PA, USA

^19^Children’s Hospital of Philadelphia, Philadelphia, PA, USA

^20^AFBS for Nemaline Myopathy (A Foundation Building Strength for Nemaline Myopathy), Palo Alto, CA, USA

^21^Department of Clinical and Biomedical Science, University of Exeter, UK

^22^Medical Genetics Branch, National Human Genome Research Institute, National Institutes of Health

^23^University of North Carolina at Chapel Hill, Chapel Hill, NC, USA

^24^Yaya Foundation for 4H Leukodystrophy, Minneapolis, MN, USA

^25^National Tay-Sachs & Allied Diseases Association, Boston, MA,USA

^26^Shwachman-Diamond Syndrome Alliance Inc, Woburn, MA, USA

^27^INADcure Foundation, Fairfield, NJ, USA

^28^Boston Children’s Hospital, Boston, MA, USA

^29^Duke University School of Medicine, Durham, NC, USA

^30^Baylor College of Medicine, Houston, TX, USA

^31^University of Florida, Gainesville, Florida, USA

^32^University of Rochester Medical Center, Rochester, NY, USA

^33^The Primary Ciliary Dyskinesia Foundation, Bloomington, MN, USA

^34^Cure LBSL, Arlington, VA, USA

^35^University of Michigan Congenital Heart Center, Ann Arbor, MI, USA

^36^Texas Children’s Hospital, Houston, TX, USA

^37^Long Island University, Brookville, NY, USA

^38^Moser Center for Leukodystrophies at Kennedy Krieger, Kennedy Krieger Institute, Baltimore, MD, USA

^39^Department of Neurology, Johns Hopkins University School of Medicine, Baltimore, MD, USA

^40^University of Pennsylvania, Philadelphia, PA, USA

^41^Congenital Hyperinsulinism International, Glen Ridge, NJ, USA

^42^McGill University Health Centre, Montreal, QC, Canada

^43^ATGC Insights, LLC, MD, USA

^44^Association for Creatine Deficiencies, Carlsbad, CA, USA

## Authors received funding as follows

Bernadette R. Gochuico: This work was funded in part by the Intramural Research Program, National Human Genome Research Institute

## Conflicts of interest are as follow

Rebecca L. Koch: Rebecca L. Koch has received grant/research support from Alnylam Pharmaceuticals, Inc.

## Notes

### Competing Interest Statement

S.M.B. was a paid consultant for Pharming Group N.V.. H.L.R. has received support from Illumina and Microsoft to support rare disease gene discovery and diagnosis. A.O.L. was a paid consultant for Addition Therapeutics and received support in the form of reagents from PacBio to support rare disease research.

### Funding Statement

Research reported in this publication was supported by by the Chan Zuckerberg Initiative Donor-Advised Fund at the Silicon Valley Community Foundation [2020-224274, 2022-316726, DAF2025-360376] (https://doi.org/10.37921/236582yuakxy) (funder DOI 10.13039/100014989).

## References

1. Kariampuzha WZ, Alyea G, Qu S, et al. Precision information extraction for rare disease epidemiology at scale. J Transl Med. 2023;21(1):157. doi:10.1186/s12967-023-04011-y

2. Guo MH, Gregg AR. Estimating yields of prenatal carrier screening and implications for design of expanded carrier screening panels. Genet Med. 2019;21(9):1940–1947. doi:10.1038/s41436-019-0472-7

3. Schrodi SJ, DeBarber A, He M, et al. Prevalence estimation for monogenic autosomal recessive diseases using population-based genetic data. Hum Genet. 2015;134(6):659–669. doi:10.1007/s00439-015-1551-8

4. Hardy GH. Mendelian proportions in a mixed population. Science. 1908;28(706):49–50. doi:10.1126/science.28.706.49

5. Hannah WB, Drumm ML, Nykamp K, Pramparo T, Steiner RD, Schrodi SJ. Using genomic databases to determine the frequency and population-based heterogeneity of autosomal recessive conditions. Genet Med Open. 2024;2(101881):101881. doi:10.1016/j.gimo.2024.101881

6. Rare As One Project. Chan Zuckerberg Initiative. January 31, 2020. Accessed February 19, 2026. https://chanzuckerberg.com/science/programs-resources/rare-as-one/

7. Chen S, Francioli LC, Goodrich JK, et al. A genomic mutational constraint map using variation in 76,156 human genomes. Nature. 2024;625(7993):92–100. doi:10.1038/s41586-023-06045-0

8. Pejaver V, Byrne AB, Feng BJ, et al. Calibration of computational tools for missense variant pathogenicity classification and ClinGen recommendations for PP3/BP4 criteria. Am J Hum Genet. 2022;109(12):2163–2177. doi:10.1016/j.ajhg.2022.10.013

9. Ioannidis NM, Rothstein JH, Pejaver V, et al. REVEL: An ensemble method for predicting the pathogenicity of rare missense variants. Am J Hum Genet. 2016;99(4):877–885. doi:10.1016/j.ajhg.2016.08.016

10. Karczewski KJ, Francioli LC, Tiao G, et al. The mutational constraint spectrum quantified from variation in 141,456 humans. Nature. 2020;581(7809):434–443. doi:10.1038/s41586-020-2308-7

11. Guez J, Goodrich JK, Moldovan MA, et al. Integrating 730,947 exome sequences with clinical literature improves gene discovery. medRxiv. Published online March 25, 2026:2026.03.23.26349081. doi:10.64898/2026.03.23.26349081

12. Singer-Berk M, Gudmundsson S, Baxter S, et al. Advanced variant classification framework reduces the false positive rate of predicted loss-of-function variants in population sequencing data. Am J Hum Genet. 2023;110(9):1496–1508. doi:10.1016/j.ajhg.2023.08.005

13. Richards S, Aziz N, Bale S, et al. Standards and guidelines for the interpretation of sequence variants: a joint consensus recommendation of the American College of Medical Genetics and Genomics and the Association for Molecular Pathology. Genet Med. 2015;17(5):405–424. doi:10.1038/gim.2015.30

14. Jee H, Huang Z, Baxter S, et al. Comprehensive analysis of ADA2 genetic variants and estimation of carrier frequency driven by a function-based approach. J Allergy Clin Immunol. 2022;149(1):379–387. doi:10.1016/j.jaci.2021.04.034

15. Riggs ER, Andersen EF, Cherry AM, et al. Technical standards for the interpretation and reporting of constitutional copy-number variants: a joint consensus recommendation of the American College of Medical Genetics and Genomics (ACMG) and the Clinical Genome Resource (ClinGen). Genet Med. 2020;22(2):245–257. doi:10.1038/s41436-019-0686-8

16. orpha.net W, orphadata.org W. Prevalence, incidence and number of published cases or families. Accessed March 30, 2026. https://www.orpha.net/orphacom/cahiers/docs/GB/Epidemiology_in_Orphanet_R1_Ann_Epi_EP_05.pdf

17. Flanagan SE, Xie W, Caswell R, et al. Next-generation sequencing reveals deep intronic cryptic ABCC8 and HADH splicing founder mutations causing hyperinsulinism by pseudoexon activation. Am J Hum Genet. 2013;92(1):131–136. doi:10.1016/j.ajhg.2012.11.017

18. Koch RL, Akman HO, Chown E, et al. Unifying the communities of early-onset glycogen storage disease type IV and adult polyglucosan body disease through a genetic prevalence study of GBE1-related disease. medRxiv. Published online December 17, 2025:2025.12.16.25342386. doi:10.64898/2025.12.16.25342386

19. Kurtovic-Kozaric A, Singer-Berk M, Wood J, et al. An estimation of global genetic prevalence of PLA2G6-associated neurodegeneration. Orphanet J Rare Dis. 2024;19(1):388. doi:10.1186/s13023-024-03275-x

20. Miyake CY, Lay EJ, Soler-Alfonso C, et al. Natural history of TANGO2 deficiency disorder: Baseline assessment of 73 patients. Genet Med. 2023;25(4):100352. doi:10.1016/j.gim.2022.11.020

21. Ghosh R, Harrison SM, Rehm HL, Plon SE, Biesecker LG, ClinGen Sequence Variant Interpretation Working Group. Updated recommendation for the benign stand-alone ACMG/AMP criterion. Hum Mutat. 2018;39(11):1525–1530. doi:10.1002/humu.23642

22. López-Rivera JA, Pérez-Palma E, Symonds J, et al. A catalogue of new incidence estimates of monogenic neurodevelopmental disorders caused by de novo variants. Brain. 2020;143(4):1099–1105. doi:10.1093/brain/awaa051

23. Gillentine MA, Wang T, Eichler EE. Estimating the prevalence of DE Novo monogenic neurodevelopmental disorders from large cohort studies. Biomedicines. 2022;10(11):2865. doi:10.3390/biomedicines10112865

24. Martin HC, Gardner EJ, Samocha KE, et al. The contribution of X-linked coding variation to severe developmental disorders. Nat Commun. 2021;12(1):627. doi:10.1038/s41467-020-20852-3

