## Supplemental Tips and Case Studies for "The Power of Partnership: Democratizing Genetic Prevalence to Empower Patient Advocacy"

### Tips for Interpretation

1. **Lack of Representation**
   - ClinGen Sequence Variant Interpretation Working Group has recommended only using allele frequencies when there are more than 2,000 reference alleles.[1]
   - All genetic ancestry-group-specific estimates should be further investigated to assess the relative representation of that group in gnomAD. Caution is advised when interpreting results for underrepresented genetic ancestry groups.
   - If a condition is known to be more common in a specific group, investigators should specifically evaluate the representation of that population in the database used for AFs.

2. **Curation of Variation**
   - Recommend performing curation for any variant with an AC≥15 (AF ≲ 1.0E-05) to ensure there is sufficient evidence of pathogenicity for the disease of interest.
     - For very rare diseases with few or no variants with AC≥15, we recommend curating at least the top 10 highest frequency variants in the variant list.
   - At a minimum, investigators should curate any purported P/LP or pLoF variant with AF higher than that of the most common, well-established LP/P variant from ClinVar.
     - GeniE provides a flag ("High AF") for these variants in order to help with identifying high-priority variants for review.
   - Predicted loss-of-function (pLoF) variants should be reviewed for flags indicating they may not result in nonsense-mediated decay.[2]

3. **Clinical Sensitivity**
   - Use results with caution for diseases where exome or genome sequencing does not yield a diagnosis in a significant portion of patients.
   - Pay attention to the allele numbers (AN; denominator of AFs) in technically challenging regions, or in genome-only regions (e.g., deep intronic variants), as the low AN may inflate the AF, especially in ancestry groups that do not have many genome samples.

4. **Gene-Disease Novelty**
   - Results from genes with newer or less established (limited/moderate) gene-disease relationships should be viewed as preliminary estimates, as the spectrum of pathogenic variants is likely not yet characterized.
   - Review the number of pathogenic and likely pathogenic variants that are available in ClinVar and other open source databases. If very few variants are known to cause disease, this process should be repeated as more variants are identified.

- This is distinct from conditions where only a small number of pathogenic variants exist (such as for some conditions with gain-of-function mechanism), though recessive conditions are generally loss of function.

5. **Disease spectrum**
    - Symptomatic carriers: Depletion of symptomatic carriers could lower the allele frequencies of pathogenic variants observed in gnomAD due to the types of cohorts that are included in the database.
    - Impacts to life expectancy: The contribution of rare genetic disease to miscarriage is not well understood for individual conditions. Those impacted pregnancies would be accounted for in these estimates, in addition to liveborn affected individuals.
    - Reduced penetrance and variable expressivity: This method assumes all variant combinations lead to the phenotype/disease of interest. Diseases where the phenotype varies based on genotype combination would all be counted together.

### Example Case Studies

#### 1. Low or limited representation of a genetic ancestry group causing an underestimate

Case Example: *HADH*

In our study, *HADH* had one of the lowest prevalences of all the groups with an estimated genetic prevalence of less than 1 in 565 million. Yet, from working with Congenital Hyperinsulinism International, we understood that this was an underestimate based on current case registry numbers. We already knew from a literature review that the Turkish founder variant (c.636+471G>T) was missing from the gnomAD dataset and that would likely influence the results. However, through personal communication with their disease experts, we realized many of the pathogenic variants in their internal database, which were not present in gnomAD, had been identified in individuals of Middle Eastern descent, a population known to be underrepresented in gnomAD. This is a clear indication that the lack of representation in population databases hinders our ability to accurately assess genetic prevalence.

2. High frequency variants that are called pathogenic by multiple laboratories but have no cases reported in the literature.

<u>Case Example: *SELENON*</u>

The c.402_403+2delGAGT variant in *SELENON* (1-26128605-CTGAG-C) is reported as pathogenic or likely pathogenic by 6 labs in ClinVar. However, in a review of the evidence provided by the labs, none of them mentioned seeing the variant in an affected individual. Additionally, after discussion with the Cure CMD organization and their scientific network, no affected cases were identified. While the allele frequency of this variant (AF = 0.0002) is not high enough to provide evidence against pathogenicity, it is the variant with the second highest allele frequency in our *SELENON* analysis. If this variant was in fact disease-causing, we would expect to see it in some affected individuals, yet there are currently no cases reported. This variant is also located in exon 3, which is included in the MANE transcript but has low per-base expression (pext) data[3], supporting the argument that this is not a biologically relevant exon. For the purposes of this study, we removed this variant despite an LP/P classification from multiple laboratories.

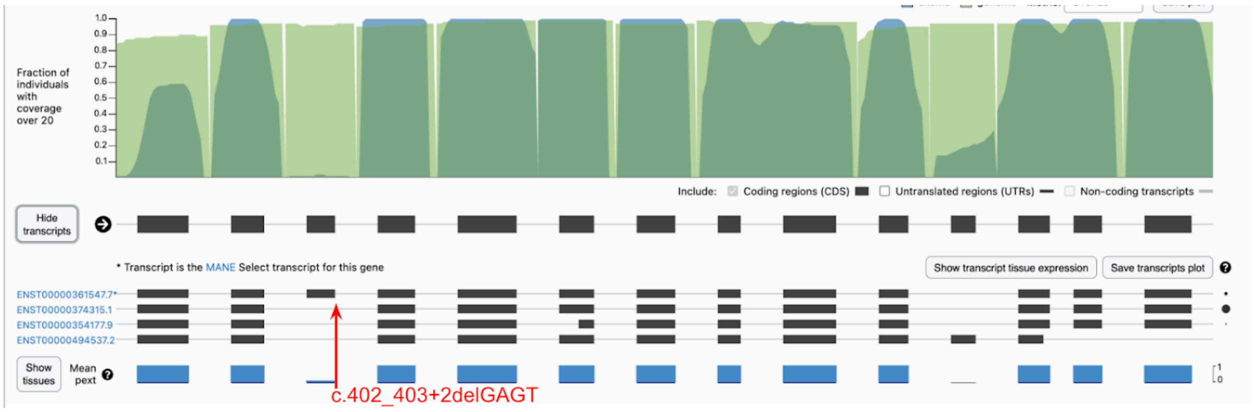


3. Hypomorphic variants that do not cause disease in homozygosity

<u>*Case Example: POLR3A*</u>

During our v2 analysis of *POLR3A*, the c.1909+22C>T (10-78009515-C-T) variant was found to be the most common disease-causing variant in the gene. However, only compound heterozygous cases have been identified to date. It was notable that these cases were all adult-onset myopathy cases, which has a very different presentation from the early-onset leukodystrophy phenotype that is primarily associated with the *POLR3A* gene. This supported the possibility of a larger phenotype spectrum, but there was no evidence to support or refute whether homozygous c.1909+22C>T individuals will or won't present with disease. This variant is likely a reduced penetrance and/or hypomorphic variant. When gnomAD v4 was launched in November 2023, there were now 10 homozygous individuals in v4, only 1 of which was in the non-UK Biobank subset. While future research could involve looking into the phenotype of homozygous cases in UK Biobank, for the purpose of this study, we removed the genetic prevalence of homozygous c.1909+22C>T cases from the final conservative and relaxed genetic prevalence estimates.

#### 4. Newer gene-disease association with high burden of missense variants

*Case Example: SLC13A5*

The gene *SLC13A5* is one of the newer gene-disease relationships in this study, associated with a non-specific phenotype of developmental delay and epileptic encephalopathy. This gene also had the second fewest number of LP/P/DM variants from ClinVar and HGMD. Additionally, the most common pathogenic variants are missense variants, which are some of the most challenging types of variants to have enough evidence to prove pathogenicity.[4] Unlike the LoF variants, which can be easier to filter and include in this analysis, figuring out which of the 299 ClinVar missense VUS in gnomAD to include in our analysis is a challenge, especially since none of the missense variants present in gnomAD had a high REVEL score (>0.932). In a recent publication, deep mutational scanning was used to reveal the effect of 90% of all possible missense variants on the structure and function of *SLC13A5* (PMID: 40577459). This method could replace other in silico tools, such as REVEL, in prioritizing missense variants to include in prevalence estimates based on the likelihood that they may be pathogenic.

#### 5. Variants with Reduced Penetrance

While we did not come across any reduced penetrance variants as a part of this study, these variants have been documented in the literature and been seen in cases. Our suggestion for how to handle these variants is to multiply the variant's AF by the approximate percentage of penetrance, and to use the new modified AF when calculating genetic prevalence. Carrier frequency estimates would remain the same.

#### 6. Breaking down numbers by phenotypic spectrum

*Case Example: GBE1 and PLA2G6*

Two genes in this study, *GBE1* and *PLA2G6,* are each associated with multiple diseases that are believed to be a part of a broader gene-disease spectrum. In *GBE1* disease-causing variants are associated with either Adult Polyglucosan Body Disease (APBD), early-onset forms of Glycogen Storage Disease Type IV or both diseases.[5] For *PLA2G6,* variants have been associated with Infantile neuroaxonal dystrophy (INAD), Atypical neuroaxonal dystrophy (atypical NAD), PLA2G6-related dystonia-parkinsonism, or in some cases multiple diseases (typically atypical NAD and parkinsonian phenotype).[6]

Not every variant in these genes have been seen in patients, which means we were not always able to associate every variant to a gene. However, we were able to group most of the variants, especially the more common ones, enabling us to provide a breakdown of the carrier frequency by disease type. We also attempted to provide a breakdown of the genetic prevalence by disease for both of these groups, however the disease outcome is not known for every variant combination and thus this should be done for informational purposes only.

#### 7. Genes with multiple inheritance patterns

*Case Example: ABCC8*

The *ABCC8* gene accounts for 40-45% of all identified cases of congenital hyperinsulinism (CHI).[7] However, the typical process for estimating AR genetic prevalence was complicated by the fact that *ABCC8* has a variable phenotypic spectrum, multiple modes of inheritance, and cases of focal CHI caused by somatic variation. To assess whether it is possible to accurately estimate the genetic prevalence of recessive disease in a gene with multiple inheritance patterns and phenotypes, we aimed to isolate the autosomal recessive (AR) CHI-only variants in *ABCC8*. However, this process proved to be challenging with some variants being associated with multiple phenotypes and inheritance patterns. Due to our conservative approach, only including variants with well-established AR CHI association, it is likely that we have underestimated the true genetic prevalence of ABCC8-associated AR CHI.

#### 8. Removing variant combinations known to be incompatible with life or not cause disease

Case Example: *DARS2*

In some diseases, not all variant combinations are known to be compatible with life and/or are disease-causing. This can be supported by certain common variants never being seen in homozygosity (as discussed above) or certain compound heterozygous combinations never being seen. This is the case with *DARS2*, where being homozygous or compound heterozygous for variants that result in no protein has not been observed in individuals with Leukoencephalopathy with brain stem and spinal cord involvement and lactate elevation (LBSL). While the literature states that nearly all patients have been found to have an exon 3 skipping variant on one allele and either a missense, nonsense, deletion or splice site variant on the other allele.[8] Personal communication with Cure LBSL and their scientific experts revealed that this is not the breakdown they see in their database.

Based on the Cure LBSL database, and what their experts are seeing in clinic, the current hypothesis is that homozygous or compound heterozygous null variants (any variant resulting in nonsense mediated decay (NMD)) are incompatible with life. Conversely the presence of at least one "leaky" variant, where at least some protein is produced, would result in some protein being present, and thus would result in an affected individual.

While the method for calculating carrier frequencies stayed the same ($2AF_a$), we adjusted our method for calculating genetic prevalence to address the known variant combinations. In order to do this, we sorted the variants into two categories, "NMD" and "non-NMD". "NMD" variants were defined as nonsense and frameshift variants where LoF curation is LoF, likely LoF, or uncertain, or variants where there was demonstrable evidence of NMD. "non-NMD" variants were all other variants where there was no evidence of NMD. We then calculated the genetic prevalence using the following formula:

$$q_a^2 - q_{nmd}^2$$

Additional research and rigorous testing of NMD would help refine these estimates over time.

#### 9. Dual Diagnoses - Large Chromosomal abnormality plus recessive condition

<u>Case Example: *TANGO2 deficiency within the 22q11.2 deletion syndrome community*</u>

*TANGO2* falls within the 22q11 region, meaning individuals with 22q11.2 deletion syndrome (DS) are at risk for *TANGO2* deficiency if they also inherited a pathogenic variant from one of their parents. Using the conservative carrier frequency of *TANGO2* (1/614), and the reported prevalence of 22q11.2DS (1/6,000),[9] we can estimate that prevalence of *TANGO2* deficiency and 22q11.2DS is:

$$(1/614 \div 2) * (1/6{,}000) = 1/7{,}368{,}000$$

We found a higher *TANGO2* carrier frequency in the Admixed American group and 22q11.2DS has a reported higher prevalence of 1:3,800 in individuals with Hispanic ethnicity. This could be an underdiagnosed dual diagnosis in this population, which warrants additional research.

The 22q11.2 deletion is observed within gnomAD (https://gnomad.broadinstitute.org/variant/GD_22Q11.2-AB__DEL?dataset=gnomad_sv_r4), with an AF of 0.00007167, or ~1/14,000, which is not surprising given the variable presentation of 22q11.2DS. However, since gnomAD excludes cohorts recruited for rare disease, we felt using gnomADs AF would have been an under estimate of prevalence and chose to use the reported prevalence instead.

#### References


1. Ghosh R, Harrison SM, Rehm HL, Plon SE, Biesecker LG, ClinGen Sequence Variant Interpretation Working Group. Updated recommendation for the benign stand-alone ACMG/AMP criterion. *Hum Mutat*. 2018;39(11):1525-1530. doi:10.1002/humu.23642

2. Singer-Berk M, Gudmundsson S, Baxter S, et al. Advanced variant classification framework reduces the false positive rate of predicted loss-of-function variants in population sequencing data. *Am J Hum Genet*. 2023;110(9):1496-1508. doi:10.1016/j.ajhg.2023.08.005

3. Cummings BB, Karczewski KJ, Kosmicki JA, et al. Transcript expression-aware annotation improves rare variant interpretation. *Nature*. 2020;581(7809):452-458. doi:10.1038/s41586-020-2329-2

4. Jaramillo-Martinez V, Sennoune SR, Tikhonova EB, Karamyshev AL, Ganapathy V, Urbatsch IL. Molecular phenotypes segregate missense mutations in SLC13A5 Epilepsy. *J Mol Biol*. 2024;436(22):168820. doi:10.1016/j.jmb.2024.168820

5. Koch RL, Soler-Alfonso C, Kiely BT, et al. Diagnosis and management of glycogen storage disease type IV, including adult polyglucosan body disease: A clinical practice resource. *Mol Genet Metab*. 2023;138(3):107525. doi:10.1016/j.ymgme.2023.107525

6. Hanna Al-Shaikh R, Milanowski LM, Holla VV, et al. PLA2G6-associated neurodegeneration in four different populations-case series and literature review. *Parkinsonism Relat Disord*. 2022;101:66-74. doi:10.1016/j.parkreldis.2022.06.016

7. De Franco E, Saint-Martin C, Brusgaard K, et al. Update of variants identified in the pancreatic β-cell KATP channel genes KCNJ11 and ABCC8 in individuals with congenital hyperinsulinism and diabetes. *Hum Mutat*. 2020;41(5):884-905. doi:10.1002/humu.23995

8. Guang S, O’Brien BM, Fine AS, Ying M, Fatemi A, Nemeth CL. Mutations in DARS2 result in global dysregulation of mRNA metabolism and splicing. *Sci Rep*. 2023;13(1):13042. doi:10.1038/s41598-023-40107-7

9. McDonald-McGinn DM, Hain HS, Emanuel BS, Zackai EH. 22q11.2 deletion syndrome. In: *GeneReviews(®)*. University of Washington, Seattle; 1993. https://www.ncbi.nlm.nih.gov/pubmed/20301696
